# Use of heated tobacco products may be associated with hypertensive disorders of pregnancy and low birth weight in Japan: An analysis of the JACSIS study

**DOI:** 10.1101/2021.04.12.21255292

**Authors:** Masayoshi Zaitsu, Yoshihiko Hosokawa, Sumiyo Okawa, Ai Hori, Gen Kobashi, Takahiro Tabuchi

## Abstract

**Background:** Little is known about heated tobacco product (HTP) use in pregnant women and associated maternal and neonatal risks for hypertensive disorders of pregnancy (HDP) and low birth weight (LBW). Thus, this study aimed to assess the status of HTP use among pregnant women in Japan and explore the risk of HDP and LBW associated with HTP use.

**Methods:** Using data from the Japan “COVID-19 and Society” Internet Survey (JACSIS) study, a web-based nationwide survey, we investigated 558 post-delivery and 365 currently pregnant women in October 2020. We assessed the prevalence of ever HTP smokers (defined as ever experiencing HTP use) in post-delivery and currently pregnant women. Among post-delivery women, we collected the information regarding HDP and LBW based on their Maternal and Child Health Handbooks (maternal and newborn records). In the multivariable regression analysis, we estimated the adjusted odds ratios (ORs) and 95% confidence intervals (CIs) of ever HTP smokers for HDP and LBW compared with those of never HTP smokers using logistic regression. A stratified analysis with respect to combustible cigarette smoking (never/ever) was also performed.

**Results:** The prevalence of ever HTP use were 11.7% and 12.6% in post-delivery and currently pregnant women, respectively. Among post-delivery women, ever HTP smokers had higher HDP incidence (13.8% vs. 6.5%, *P*=0.03), with an OR of 2.78 (95% CI 0.84–9.15) and higher LBW incidence (18.5% versus 8.9%, *P*=0.02), with an elevated OR of 2.08 (95% CI 0.80–5.39). A similar tendency was observed among never and ever combustible cigarette smokers.

**Conclusion:** In Japan, the incidence of HTP use has exceeded 10% among pregnant women, and HTP smoking may be associated with increased maternal and neonatal risks. School-based tobacco prevention and cessation programs should be conducted regardless of product types to prevent life-threatening perinatal complications and deaths.

**What this paper adds:**

- Little is known about heated tobacco product (HTP) use and associated perinatal risks among pregnant women.
- In Japan, the prevalence of ever HTP use exceeded 10% among pregnant women.
- HTP use approximately doubled perinatal risk of hypertensive disorders of pregnancy and low birth weight.
- When stratified by cigarette smoking status, a similar tendency was observed among never and ever cigarette smokers.

## INTRODUCTION

The widespread use of heated tobacco products (HTPs) is an emerging public health concern.[1] Since the initial marketing of HTPs in 2014, the prevalence of HTP use has increased in Japan, exceeding 15% in the young population aged 20–39 years in 2019,[2] and this incidence was maintained over 15% during the coronavirus disease (COVID-19) pandemic in 2020.[3]

Although the impression of HTPs as a healthy alternative to combustible cigarettes is promoted by the advertising of HTPs (e.g., reduced harmfulness and a smoke-free image),[4] HTP-related unfavorable health outcomes, including acute respiratory and cardiovascular risks, are likely to occur.[5, 6] However, little is known about HTP use and associated maternal and neonatal risks in pregnant women, including hypertensive disorders of pregnancy (HDP) and low birth weight (LBW).[7, 8] Although some controversial associations have been reported for HDP with respect to combustible cigarettes,[9] this type of cigarettes increases various maternal and neonatal risks in Japan.[10, 11] Therefore, we hypothesized that HTP use is associated with HDP and LBW, regardless of combustible cigarette smoking.

This study aimed to assess the status of HTP use among pregnant women in Japan and explore the risk of HDP and LBW associated with the use of HTP by analyzing data from a nationwide web-based survey in Japan that contained pregnancy-related information and data related to behavioral factors (e.g., HTP use and combustible cigarette smoking), and social background.

## MATERIALS AND METHODS

### Data setting

This cross-sectional internet-based study is part of the Japan COVID-19 and Society Internet Survey (JACSIS) study. The JACSIS study comprises three surveys in the following three target populations: (a) young people and adults aged 15–79 years, (b) currently pregnant and post-delivery women, and (c) adults living in a single-parent household. The study samples for each survey were retrieved from the pooled panels of an internet research agency (Rakuten Insight, Inc., which had approximately 2.2 million panelists in 2019).[12] We used data from currently pregnant and post-delivery women, which were collected in October 2020.

The internet research agency identified 21,896 eligible women, randomly selected 4373 women who gave birth after October 2019 or who were expected to give birth by March 2021, and distributed the questionnaire comprising 61 questions to the selected women through a designated website. Next, we collected data from 1000 women (response rate, 22.9%) stratified by delivery date as follows: (a) 600 post-delivery women who delivered during October 2019–March 2020 (n=200), April–May 2020 (n=200), and June–October 2020 (n=200) and (b) 400 currently pregnant women who were expected to deliver during October 2020–March 2021. Among 1000 study participants, we excluded 77 who provided irrelevant or conflicting information (45 post-delivery and 32 currently pregnant women) as done in previous studies of the same research agency,[13] yielding a total of 923 study participants for the analysis (558 post-delivery and 365 currently pregnant women). Informed consent was obtained electronically, and the Institutional Review Board of the Osaka International Cancer Institute approved the study (Protocol Number 20084).

### Definition of HDP and LBW

Data on HDP and LBW were extracted from the web-based self-reported questionnaires. We defined the incidence of HDP based on whether the study participants had been diagnosed as having HDP or preeclampsia during pregnancy. The criteria for HDP diagnosis in Japan were derived from the criteria of the American College of Obstetricians and Gynecologists (i.e., systolic blood pressure ≥140 mmHg or diastolic blood pressure ≥90 mmHg after the 20th week of gestation).[14] We defined the incidence of LBW on the basis of the diagnosis of LBW (birth weight <2500 g).

All participants were asked to provide information from their Maternal and Child Health Handbooks. In brief, as previously described,[15, 16] the Maternal and Child Health Handbooks are well-established integrated home-based records of maternal, newborn, and child health. As a part of a national maternal and child health policy, all municipalities issue a handbook to all women who report a pregnancy, and medical professionals record the health information of the mother and child, including clinical outcomes (e.g., blood pressure and birth weight) and incident diagnoses (e.g., HDP and LBW) during pregnancy. Mothers seldom lose their Maternal and Child Health Handbooks (losing rate, <1%).[15]

### HTP and cigarette smoking and other covariates

In the questionnaire, study participants were asked to indicate their smoking status (never, once or a few times but not habitually, former, sometimes, or every day) for each HTP available in the study period (Ploom Tech, Ploom Tech plus, Ploom S, IQOS, glo, glo sens, and PULZE). If they answered “never” for all HTPs, we defined them as never HTP smokers; the remaining participants were considered ever HTP smokers.

We also classified the status of combustible cigarette smoking (never/ever). For other covariates, we included age, educational attainment (≤12 years [high school] or ≥13 years [college or university]), occupation (manager or others), household income (<2 million JPY [approximately 20,000 USD], 2 to <6 million JPY, and ≥6 million JPY), and comorbidity (having hypertension or diabetes).[17]

### Statistical analysis

Descriptive statistics were computed, and t-test or chi-squared test was performed. We assessed the prevalence of ever HTP smokers among post-delivery and currently pregnant women. Additionally, we described detailed HTP smoking status cross-classified according to the combustible cigarette smoking status of currently pregnant and post-delivery women.

To assess the potential association between HTP smoking and perinatal risk of HDP and LBW, we restricted the sample to 558 post-delivery women who could complete all the assessments during their pregnancy (Table 1). In the multivariable logistic regression analyses, the odds ratio (OR) and 95% confidence interval (CI) of ever HTP smokers for HDP risk were estimated with adjustment for age (model 1, the main model in the present study). The reference group comprised never HTP smokers. In model 2, we fully adjusted for other explanatory variables (combustible cigarette smoking, educational attainment, occupation, household income, and comorbidity) and excluded 64 participants with missing information on household income. The same analyses were performed for LBW. For sensitivity analysis, we conducted a stratified analysis with respect to combustible cigarette smoking (never/ever).

**Table 1.**
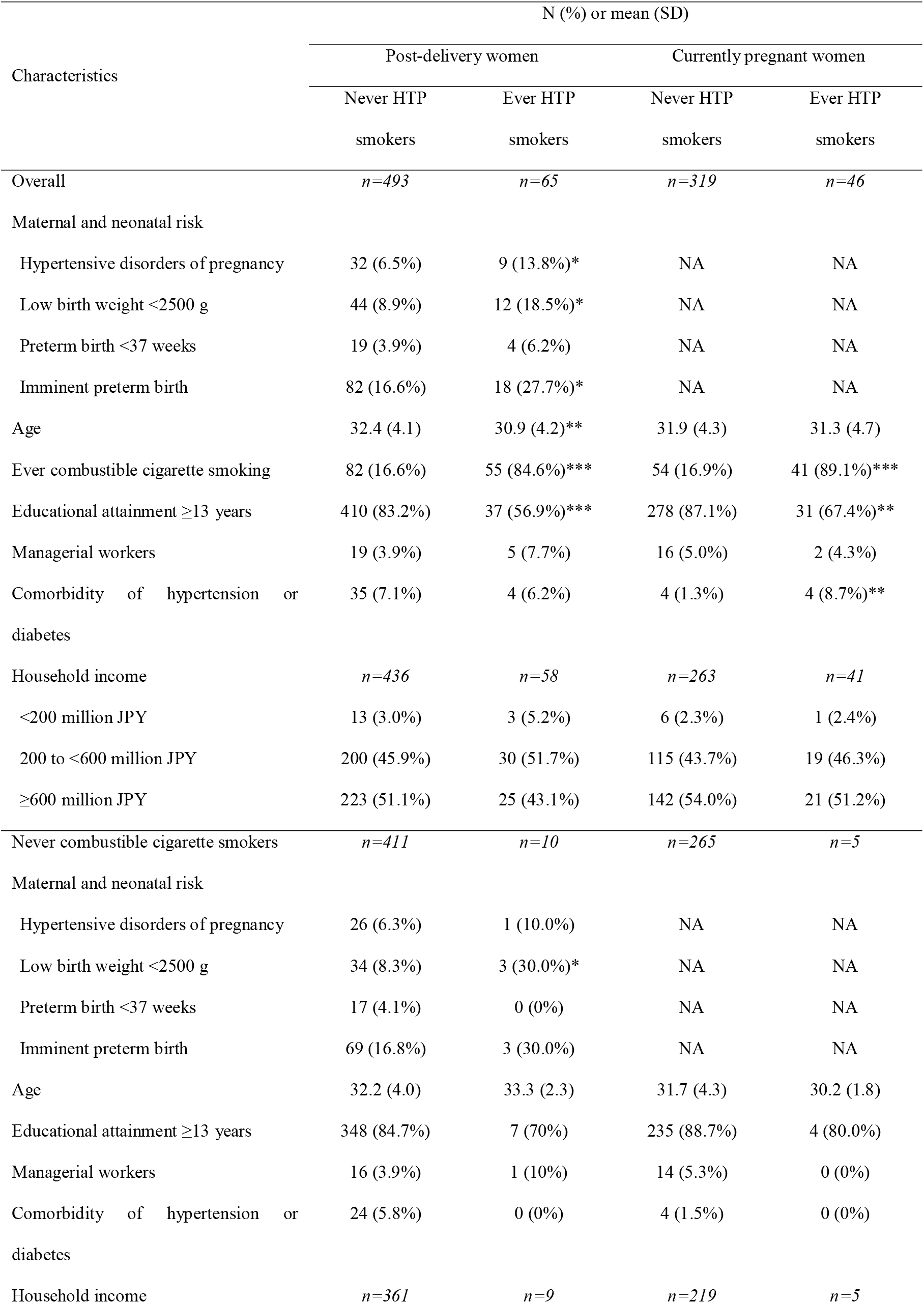
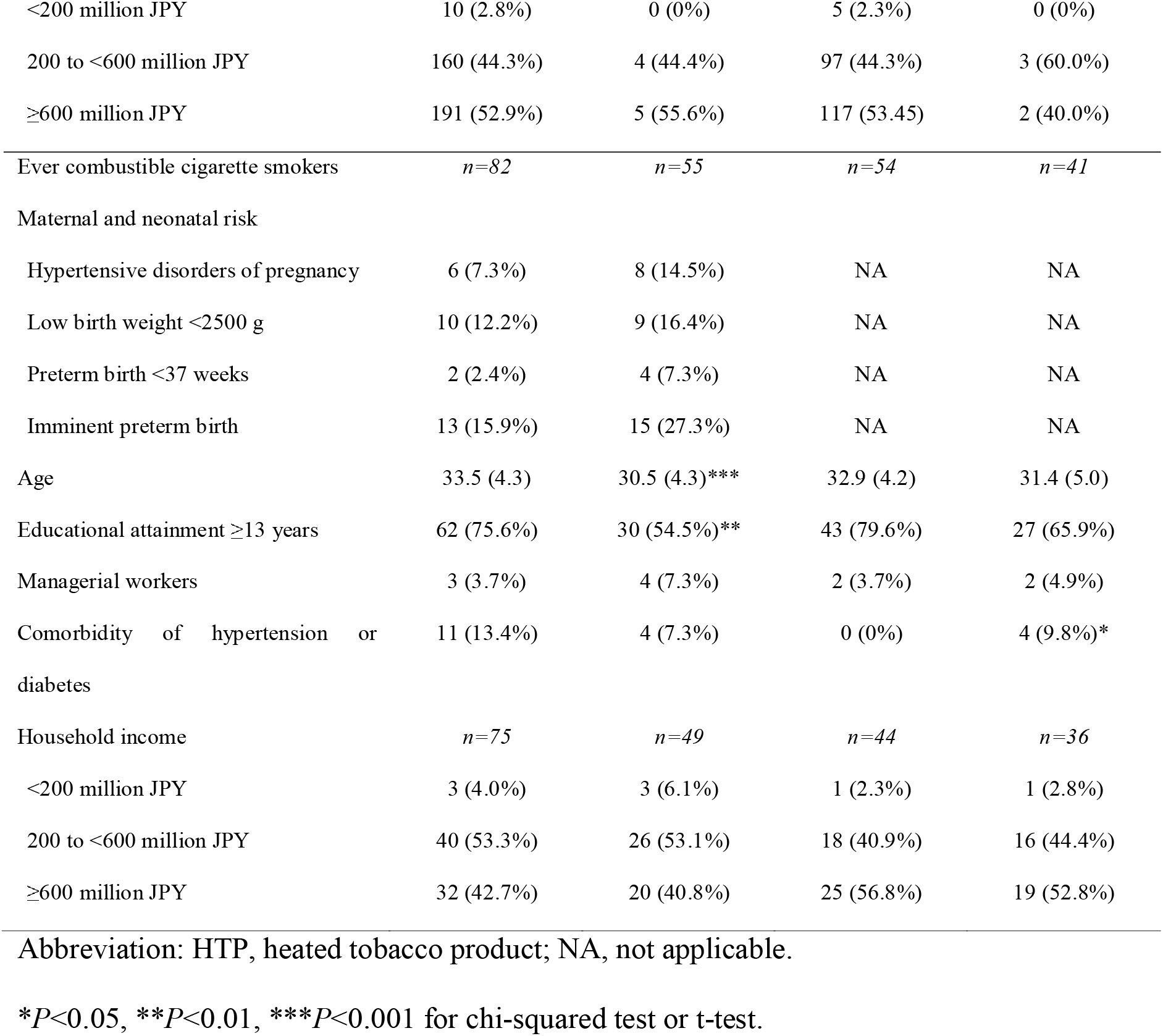
Characteristics of 558 post-delivery women and 365 currently pregnant women.

Alpha was set at 0.05, and all *P*-values were two sided. Data were analyzed using STATA/MP13.1 (StataCorp LLC, College Station, TX).

## RESULTS

Among 558 post-delivery women, the incidences of HDP and LBW were 7.3% (n=41) and 10.0% (n=56), respectively, and the prevalence of ever HTP smokers was 11.7% (n=65, Table 1). Furthermore, among 365 currently pregnant women, the prevalence of ever HTP smokers was 12.6% (n=46), which did not differ from that of HTP smokers among post-delivery women (*P*=0.66). Among currently pregnant women, 4.4% of former combustible cigarette smokers reported smoking HTPs during pregnancy (Table 2), corresponding to 1.1% (4 out of 365) of current HTP smokers.

**Table 2.** **Detailed smoking status and use of heated tobacco products cross-classified according to combustible cigarette smoking status**

| Characteristics | HTP smoking status |  |  |  |
| --- | --- | --- | --- | --- |
|  | Never | Former | Current | Total |
| Post-delivery women, $n=558$ | | | | |
| Never combustible cigarette smokers | 411 (97.6%) | 9 (2.1%) | 1 (0.2%) | 421 (100%) |
| Former combustible cigarette smokers | 79 (64.2%) | 32 (26.0%) | 12 (9.8%) | 123 (100%) |
| Current combustible cigarette smokers | 3 (21.4%) | 9 (64.3%) | 2 (14.3%) | 14 (100%) |
| Currently pregnant women, $n=365$ | | | | |
| Never combustible cigarette smokers | 265 (98.1%) | 5 (1.9%) | 0 (0%) | 270 (100%) |
| Former combustible cigarette smokers | 54 (59.3%) | 33 (36.3%) | 4 (4.4%) | 91 (100%) |
| Current combustible cigarette smokers | 0 (0%) | 4 (100%) | 0 (0%) | 4 (100%) |

Abbreviation: HTP, heated tobacco product.
\* $P<0.05$ , \*\* $P<0.01$ , \*\*\* $P<0.001$ for chi-squared test or t-test.

Among post-delivery women, the HDP incidence was higher in ever HTP smokers than in never HTP smokers (13.8% vs. 6.5%; Table 1). Similarly, the incidence of LBW was higher among ever HTP smokers than among never HTP smokers (18.5% vs. 8.9%, Table 1). When stratified by combustible cigarette smoking, a similar tendency was observed among never and ever combustible cigarette smokers (Table 1).

In the regression analysis, the age-adjusted ORs for HDP and LBW were elevated in ever HTP smokers (model 1, Figure 1); the ORs for HDP and LBW were 2.48 (95% CI, 1.11–5.53) and 2.36 (95% CI, 1.16–4.78), respectively. Although the elevated ORs were attenuated after fully controlling for other covariates, the tendency remained elevated (model 2, Figure 1). In the same regression analyses (model 2), while ever combustible cigarette smokers did not predict perinatal outcomes, managerial workers predicted the incidence of HDP and LBW; the ORs for HDP and LBW were 3.92 (95% CI 1.16–13.2) and 3.74 (95% CI 1.41–9.93), respectively. When stratified by combustible cigarette smoking, a similar tendency was observed independently in never and ever combustible cigarette smokers (Figure 1). For instance, among never combustible cigarette smokers, the age-adjusted OR of HTP use for LBW was 4.82 (95% CI, 1.19–19.6).

**Figure 1.**
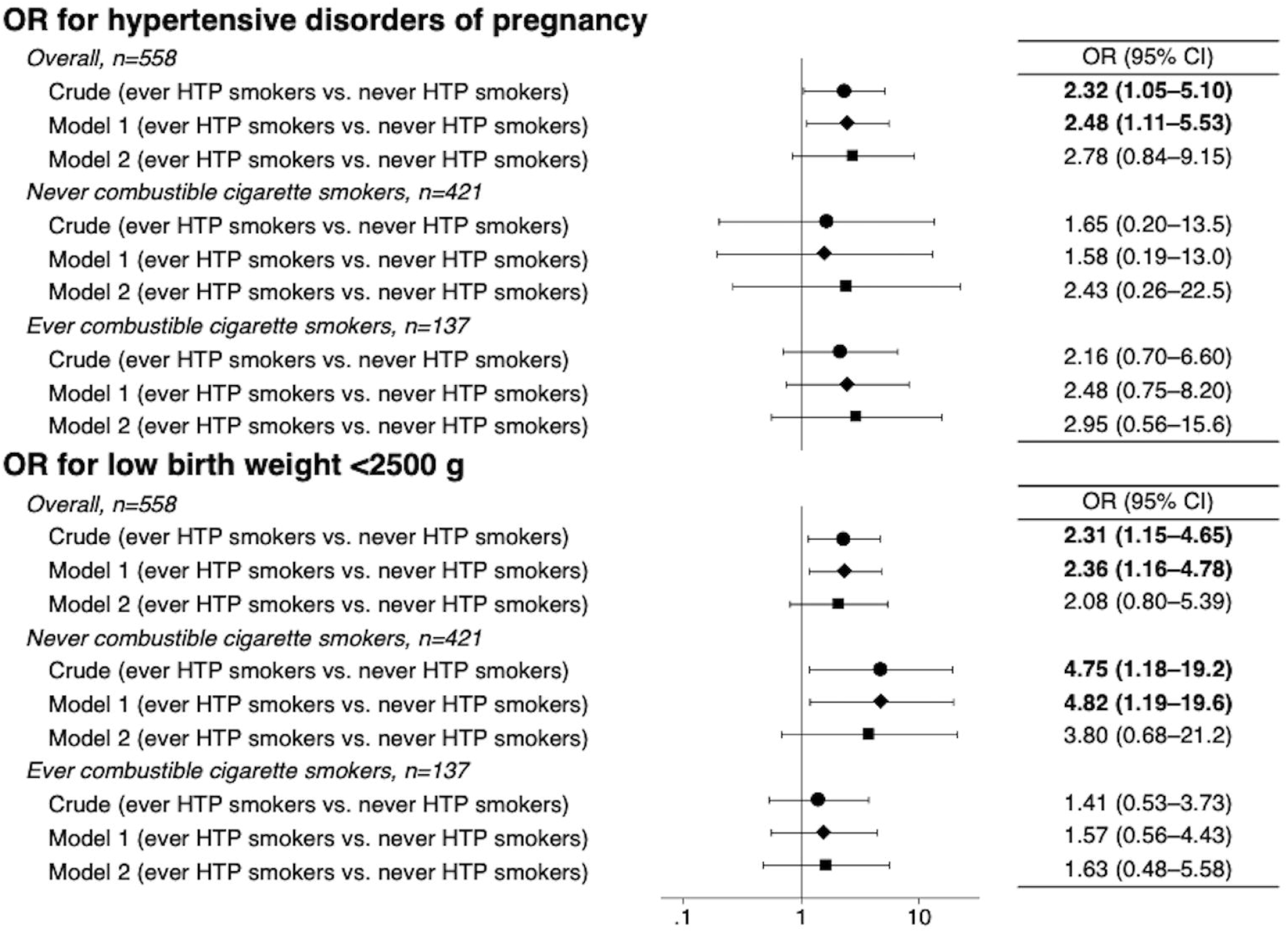
Odds ratio of ever heated tobacco product smokers with hypertensive disorders of pregnancy and low birth weight compared with never heated tobacco product smokers. Age was adjusted in model 1, and other covariates (combustible cigarette smoking, educational attainment, occupation, household income, and comorbidity) were additionally adjusted in model 2. The samples for each analysis in model 2 were as follows: n=494 (overall), n=370 (never combustible cigarette smokers), and n=124 (ever combustible cigarette smokers) for hypertensive disorders of pregnancy; and n=478 (overall), n=310 (never combustible cigarette smokers), and n=118 (ever combustible cigarette smokers) for low birth weight. Abbreviations: HTP, heated tobacco products; OR, odds ratio.

## DISCUSSION

During the COVID-19 pandemic in Japan, the incidence of HTP use among pregnant women is likely to exceed 10%, and we found that HTP use may be associated with perinatal risk of HDP and LBW. Although the impact was attenuated after controlling for other potential explanatory factors and the significance disappeared due to weak statistical power, the maternal risk might be high independent of combustible cigarette smoking. This result seems to be reliable because the incidence estimate of approximately 7% HDP found in our study (using Maternal and Child Health Handbooks) is consistent with the general statistics reported for Japanese pregnant women.[14] In addition, pregnant women of high socioeconomic status independently predicted the risk of HDP, which might also support our findings because they are known to use HTP more frequently than women of lower socioeconomic status.[17]

We also found that LBW, a well-known smoking-related neonatal risk,[18] was associated with HTP use. In fact, the incidence of HTP use doubled the risk of LBW, and the association was stronger among never combustible cigarette smokers. These results seem to be reliable because the incidence estimate of approximately 10% LBW found in our study (using Maternal and Child Health Handbooks) is consistent with the general statistics reported for Japanese pregnant women.[19] This also implies that aerosols of HTPs containing nicotine and other inhalable substances can cause acute adverse health events on the development of infants.

To the best of our knowledge, this is the first report of a potential association between HTP use and perinatal risks. Although smoking plays a controversial role,[9] recent evidence suggests that combustible cigarette smoking is associated with increased HDP risk.[10, 11] Another study also reported the risk of snuff use for preeclampsia, a severe phenotype of HDP.[20] Although the biological and genetic pathways (e.g., CYP2A6 and nicotine) underlying the associations observed in different phenotypes of HDP (e.g., preeclampsia and gestational hypertension) have not been elucidated,[21] HDP is recognized as a systemic disease attributable to placental circulatory dysfunction.[22] In experimental research, aerosol from HTPs was found to damage vascular endothelial function in rats.[5] Therefore, HDP risk associated with HTP use may involve acute and chronic vascular damage, irrespective of combustible cigarette smoking. Furthermore, as concluded in a recent systematic review, smoking is a strong risk factor for LBW.[18] Thus, given the fact that HTPs are smoking devices, our observed results are in line with established knowledge.

Finally, the impression of HTPs as a healthy alternative is promoted by the advertising of HTPs.[4] Indeed, among currently pregnant women, approximately 4% of former combustible cigarette smokers reported smoking HTPs in the present study. This result might reflect a change from combustible cigarettes to HTP smoking during pregnancy. However, our findings imply that HTP use is at least not a healthy alternative. Evidence for unfavorable health outcomes regarding HTPs is still lacking, particularly in the young population of reproductive age. Insufficient health knowledge may have led to the current increase of HTP use among pregnant women, as reflected in our results and the latest statistics in Japan.[2, 3] However, the question remains as to how multidimensional factors of the COVID-19 pandemic (e.g., the infection, mental health, and socioeconomic factors) and the smoking behaviors of others (e.g., partners and family) affect the association between HTP use and perinatal risks. Our sequential series of the JACSIS study planned in 2021 may provide updates regarding the present results.

Our study had some limitations. First, our cross-sectional design does not allow to conclude causal mechanisms between HTP use and perinatal risks. However, the prevalence of HDP and HTP smokers were mostly parallel to the general population in Japan.[2, 3, 14] In addition, the incidence of HTP use did not differ between post-delivery and currently pregnant women in our study. Second, recall and reporting bias cannot be discarded. Because self-report-based smoking status among pregnant women tends to misclassify ever smokers as never smokers,[23] our estimates might be biased toward the null. Third, the perinatal clinical information was self-reported and not based on medical charts, thereby limiting the precision of the results. However, all participants were asked to base their responses on their Maternal and Child Health Handbooks, a well-established home-base maternal and neonatal record during pregnancy.[16] Therefore, this limitation might not have affected our results or at least not largely. Despite these limitations, the strengths of the study included detailed information for HTPs, which covered all HTPs available during the study period.

Additionally, this is the first report regarding the status of HTP use among pregnant women in Japan, and it highlights the potentially elevated maternal and neonatal risks associated with HTP use. Additionally, besides the present study, no other human studies to date have assessed the potential effect of the maternal use of new tobacco products (i.e., e-cigarette and HTP) on perinatal health.[24] Therefore, our findings shed light and motivate further investigations to estimate the life-threatening perinatal risks associated with new tobacco products.

In conclusion, the incidence of HTP use seems to exceed 10% among pregnant women, and HTP smoking may be associated with increased maternal and neonatal risks in Japan. With no doubt, smoking in reproductive age can cause unfavorable perinatal outcomes.[25] Hence, efforts should be made to investigate the risk of HTP use in reproductive age, and school-based tobacco prevention and cessation programs should be conducted regardless of product types to prevent life-threatening perinatal complications and deaths.

## Data Availability

The data that support the findings of this study are available on reasonable request. However, restrictions apply to the availability of these data due to personal identification; research data are not shared. If any person wishes to verify our data, they are most welcome to contact the corresponding author.

## ACKNOWLEDGMENTS

We would like to thank Editage (www.editage.com) for English language editing.

## AUTHOR CONTRIBUTIONS

Conception and design: M Zaitsu, T Tabuchi; Development of methodology: M Zaitsu, Y Hosokawa, S Okawa; Acquisition of data: S Okawa, A Hori, and T Tabuchi; Analysis and interpretation of data: M Zaitsu, Y Hosokawa; Writing, review and/or revision of the manuscript: All authors; Study supervision: T Tabuchi.

